# The effects of developmental trauma on reinforcement learning and its relationship to psychotic experiences: a behavioural study

**DOI:** 10.1101/2020.11.18.20234112

**Authors:** Rowan Rezaie, Mustapha Modaffar, Paul Jung, Chandni Hindocha, James A Bisby, Michael A P Bloomfield

## Abstract

**Background:** Developmental psychological trauma can impact several key neurocognitive domains, including reward processing, and is associated with increased risk of psychosis in adulthood. Aberrant reinforcement learning (RL), an important component of reward processing, has been implicated in the pathophysiology of psychosis by altering information processing through changes in hierarchical predictive coding. We therefore sought to investigate RL in survivors of developmental trauma and its relationship to psychotic experiences.

**Methods:** We recruited two groups of adults, one with self-reported exposure to multiple forms of developmental trauma (*n*=115), and a control group without any known history of maltreatment (*n*=85). Participants completed measures of psychotic experiences (Community Assessment of Psychic Experiences) and undertook a probabilistic selection task designed to assess RL from positive versus negative outcomes. We analysed group differences for main effects and investigated relationships between developmental trauma, RL and psychotic experiences using regression modelling and mediation analysis.

**Results:** Developmental trauma was associated with psychotic experiences (adjusted R^*2*^=0.41, *p*=0.004) and impaired RL (*F*_*df*_=6.29_1,89_, *p*=0.014). Impaired RL mediated the association between developmental trauma and psychotic experiences (indirect effect *β* = 0.60, 95% CI, 0.01–1.36).

**Conclusions:** Our findings implicate aberrant RL as a possible mechanism through which developmental trauma may increase risk of psychosis. Further research is therefore warranted to understand the specific processes that characterise these putative trauma-induced vulnerability mechanisms and how they may contribute to the development of psychopathology.

## 1. INTRODUCTION

### 1.1 Developmental trauma and psychosis

Psychosis, a major contributor to global disease burden, is a potentially devastating illness characterised by distortions in reality processing [1]. A growing body of evidence has indicated that exposure to psychologically traumatic experiences during childhood and/or adolescence, including all forms of abuse and neglect (hereafter, ‘developmental trauma’), increases risk of psychosis in adulthood [2,3,4]. Evidence fulfilling the Bradford Hill criteria including strong, temporal, and dose-response relationships, substantiates a potential causative association between developmental trauma and psychosis, accounting for approximately one-third of cases of psychosis [2,5,6,7]. Crucially, individuals experiencing psychosis with a history of developmental trauma are at higher risk of more severe illness, increased comorbidity, re-hospitalisation, and poorer response to conventional treatment [3,8]. There is therefore a pressing need to improve treatments in this population. However, the precise mechanisms underlying this association remain poorly understood, contributing to a lack of evidence-based treatments for these clinical groups [9].

### 1.2 Hierarchical predictive coding, reinforcement learning, and psychosis

As childhood and adolescence are critical periods for both brain and psychological development, trauma-induced alterations to neurocognitive systems are thought to play an influential role in the relationship between developmental trauma and psychosis. The hierarchical predictive coding (HPC) framework offers a unifying explanatory account of brain function that may explain how developmental trauma induces vulnerability for psychosis. According to this framework, prior beliefs (‘priors’) and sensory (perceptual) inputs are weighted according to their precision (inverse variance) and compared to generate a prediction error (PE) signal, which captures the difference between the expected and actual outcome. These PE signals can be sent up to higher levels of the inferential hierarchy to update internal models of the outside world [10,11,12]. Increasing evidence indicates that the balance in precision between prior beliefs and sensory inputs is altered in people with psychosis, including recent computational work suggesting that hallucinations may result from strong perceptual priors of the external environment [13].

The HPC framework underpins reinforcement learning (RL) theory, which describes how the brain processes feedback and learns from prior experience [14,15]. Within this model, aberrant RL contributes to inflexible belief updating through an uncorrected discrepancy between priors and sensory evidence which results in the formation of perceptual inferences that are not reflective of reality [16,17,18]. Emerging evidence indicates that exposure to developmental trauma may compromise the neurocomputational processes fundamental to RL [8,19,20,21]. There is also evidence that developmental trauma can lead to lasting structural and functional alterations in brain regions thought to support these cognitive processes including the striatum and prefrontal cortex [22]. Importantly, these regions are also affected in individuals with psychosis [23,24]. Taken together, these findings indicate that aberrant RL may represent a mechanism through which developmental trauma induces vulnerability to psychosis.

### 1.3 The present study

We therefore sought to investigate the relationship between RL and psychotic experiences in survivors of developmental trauma. To investigate RL, adults with and without developmental trauma histories completed a probabilistic selection task [25,26] and behavioural data was computationally modelled using a hierarchical Bayesian inference framework to assess learning from positive and negative feedback [27,28]. Given evidence of attentional biases towards negative stimuli among developmental trauma survivors [8,29], we hypothesised that the developmental trauma group would display impaired positive feedback (‘Go’) learning. In accordance with HPC accounts of psychosis [11,18], we also hypothesised that deficits in RL would, in part, account for the relationship between developmental trauma and psychotic experiences.

## 2. MATERIALS AND METHODS

This study received ethical approval from the University College London (UCL) Research Ethics Committee (14317/001). All participants provided informed consent prior to participation.

### 2.1 Participants and procedure

We recruited participants via online and social media advertising. Participant inclusion criteria were: (1) good physical health; (2) UK-based; (3) fluent in English; (4) age 18-65 years; (5) access to a computer to undergo the study; (6) ability to give informed consent. Self-reported present psychiatric diagnosis, present psychiatric medication use, and/or any past or current major medical condition led to exclusion from the study. Additional inclusion criteria based on self-reported exposure to developmental trauma is specified below. Participants completed a battery of clinical questionnaires and undertook a probabilistic selection task [25, 26] on a web-based interface, *www.gorilla.sc* [30], and were entered into a gift voucher prize draw (£50) for their participation.

### 2.2 Measures

#### 2.2.1 Developmental trauma

We assessed self-reported exposure to developmental trauma using the 25-item Childhood Trauma Questionnaire (CTQ) [31]. The CTQ is a widely used tool for developmental trauma with good psychometric properties in patients with psychosis (Cronbach’s alpha=0.89; [32]), and measures five distinct subtypes of traumatic experiences before the age of 17: emotional abuse, physical abuse, sexual abuse, emotional neglect, and physical neglect (Supplementary Methods).

For each item, participants were asked to gauge their responses on a Likert scale from 1 (never true) to 5 (very often true). Pre-defined threshold scores [31] were used to classify exposure to each trauma type (Supplementary Table S1). Inclusion criteria for the developmental trauma (DT+) group included at least moderate self-reported exposure to more than two types of trauma, and for the control (DT-) group, below moderate self-reported exposure to all five types of trauma (Supplementary Methods).

#### 2.2.2 Psychotic experiences and psychosis proneness

We assessed psychotic experiences using the 15-item Community Assessment of Psychic Experiences–Positive scale (CAPE-P15) [33]. The CAPE-P15 is a self-report questionnaire which assesses feelings of persecutory ideation, bizarre experiences, and perceptual abnormalities on a 4-point Likert scale (1 = never, 4 = nearly always) and been shown a reliable measure of recent psychotic-like experiences in the general population (Cronbach’s alpha=0.79; [33]). Ultra-high risk for psychosis status was assessed using a pre-defined mean cut-off score of 1.47 [34].

We used the abridged Oxford-Liverpool Inventory of Feelings and Experiences (sO-LIFE) [35] as a measure of psychosis proneness (i.e. schizotypy). The 43-item sO-LIFE measures four dimensions; unusual experiences, cognitive disorganisation, introvertive anhedonia, and impulsive non-conformity, and has been demonstrated psychometrically reliable (Cronbach’s alpha=0.62-0.80; [35]).

#### 2.2.3 Confounding variables

We examined potential demographic confounds including age, sex, ethnicity, educational attainment, smoking history, prior use of secondary care mental health services and/or psychiatric medication, and childhood socioeconomic status (SES) (Supplementary Methods).

We also used the Quick Inventory of Depressive Symptomatology (QIDS) [36] and the Spielberger State-Trait Anxiety Inventory (STAI) [37] to assess and control for depressive symptoms and anxiety respectively. Further assessment included screening for drug use via the Drug Abuse Screening Test (DAST-10) [38] and alcohol use via the Short Michigan Alcoholism Screening Test (SMAST) [39].

### 2.3 Probabilistic selection task

Participants completed a probabilistic selection task [25,26] consisting of two phases: a learning phase and a transfer phase (Figure 1). In the learning phase, participants were randomly presented with one of three different stimulus pairs on each trial (images of decks of cards; Figure 1) and were tasked with learning to select the most rewarded stimulus in each pair through prior choice and feedback. Participants received probabilistic feedback for winning (+1 point) or losing (0 points) choices; stimulus A was rewarded in 80% of AB pair trials while stimulus B was rewarded in the remaining 20% of AB trials; similarly 70% and 30% in CD pairs, and 60% and 40% in EF pairs (Figure 1). Participants completed three blocks of 60 trials (20 trials per stimulus pair) with the aim of scoring as many points as possible.

**Figure 1:**
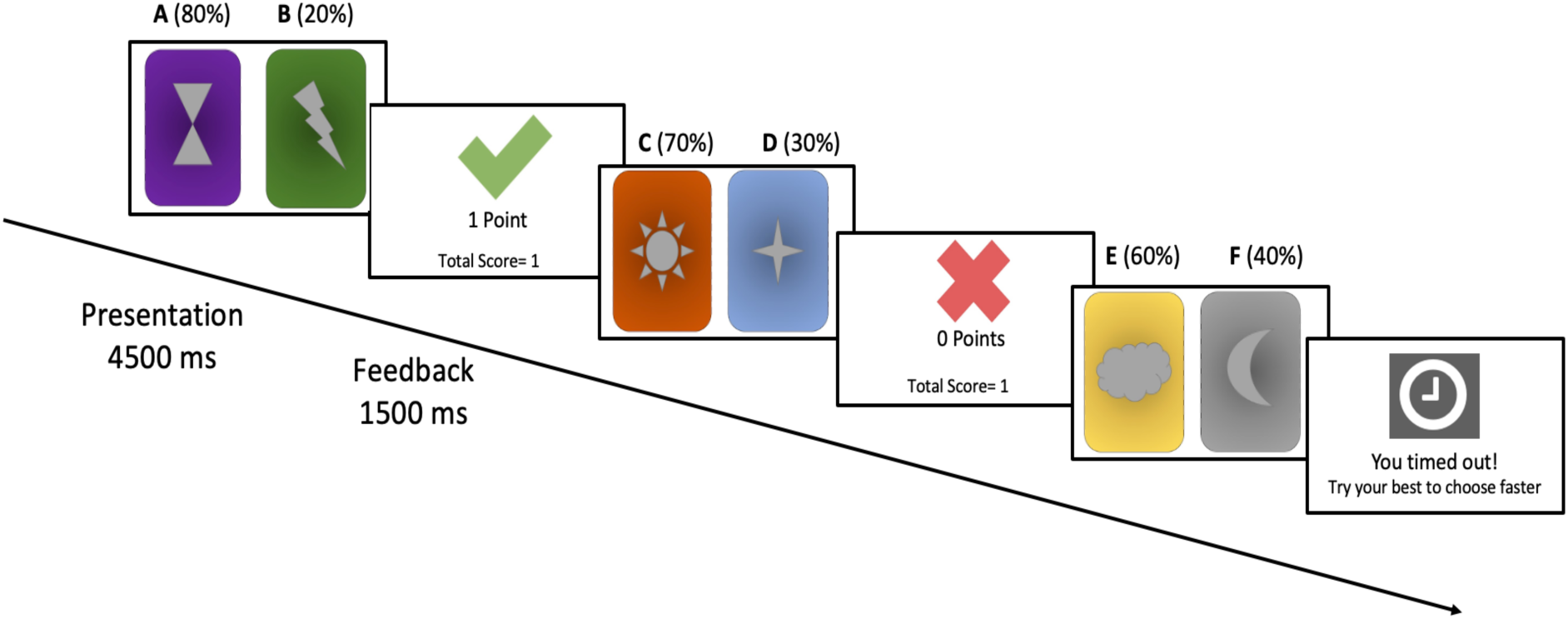
Experimental paradigm. In the learning phase (3 blocks of 60 trials), stimuli (decks of cards) were displayed for 4500ms, during which participants were asked to pick a card from one of the two decks with the aim of scoring as many points as possible. Feedback was presented for 1500ms. No feedback was provided if participants chose not to respond to a particular stimulus. In the transfer phase (32 test trials), novel stimulus combinations of either card A or card B were presented with a fixation cross displayed in place of feedback. Choosing A, the most rewarded stimulus is dependant on positive feedback-driven learning while avoiding B, the least rewarded stimulus, is indicative of negative feedback-driven learning.

In the transfer phase, novel pairings of either stimulus A (AC/AD/AE/AF) or B (BC/BD/BE/BF), were presented without feedback to assess whether learning was better accomplished via positive feedback (‘Go’ learning), as indicated by choose-A performance, or negative feedback (‘NoGo’ learning), as indicated by avoid-B performance. In addition to the 32 test trials (four trials per stimulus pair), four further AB pairs were presented to ascertain acquired learning of the most marked contingencies. Participants who had failed to identify A as the more rewarding stimulus on less than 3 of the 4 pairings were excluded from analysis in the transfer phase.

### 2.4 Computational model

We fit a hierarchical Bayesian Q-learning model [28] with learning phase behavioural data to model prediction-based learning on a trial-by-trial basis. The Q-learning gain/loss model [26,27] is a variation of the Q-learning RL algorithm [14] with three free parameters. Separate learning rate parameters for gain (*α*_*G*_), and loss (*α*_*L*_) outcomes were used to determine learning from positive versus negative feedback, and an exploration-exploitation parameter (*β*) was used to assess deterministic choice behaviour (i.e. how much participants exploited differences in stimulus contingencies). In a hierarchical Bayesian framework, individual and group parameter distributions were simultaneously fit to assess acquired learning [28,41]. Further detail of the model is presented in the Supplementary Information.

### 2.5 Statistical analyses

Statistical analyses were conducted using SPSS version 26 (IBM Corp., Armonk, N.Y.) and RStudio (version 1.2.5033). Statistical significance was referenced at *p*<0.05 (two-tailed) for all tests. Independent sample *t*-tests and *χ*_*2*_ tests were used to investigate whether potential confounding clinico-demographic variables reported to impact RL were also associated with exposure to developmental trauma in our sample.

First, we assessed associations between developmental trauma and psychotic experiences and schizotypy. Hierarchical multiple regression analyses were conducted to examine whether exposure to developmental trauma (DT+ vs DT-) significantly predicted psychotic experiences and schizotypy scores after adjusting for candidate confounds (Supplementary Methods).

Second, we examined the effects of developmental trauma on behavioural performance in the learning phase using repeated measures analyses of variance (ANOVAs) with block and reward contingency pair (AB/CD/EF) as within-subjects factors and group (DT+/DT-) as a between-subjects factor. Appropriate *post hoc* tests were conducted to compare average stimulus-specific block performance between groups. The hierarchical Bayesian Q-learning gain/loss model was implemented using the *hBayesDM* R package version 1.0.2 [41] and a confirmatory model-fit assessment was conducted (Supplementary Methods). Bayes Factor analyses and frequentist analyses (*t*-tests) were performed to compare posterior distributions of modelled group-level parameters. Statistical significance was also inferred if compared highest density intervals (HDI) did not overlap 0 [41]. Cohen’s *d* effect sizes were calculated to characterise inter-group differences in behavioural measures and Pearson correlation analyses were performed to assess relationships between task performance, psychotic experiences and schizotypy.

Finally, we conducted mediation analyses to investigate whether RL mediated the effect of developmental trauma on psychotic experiences using the Hayes PROCESS macro for SPSS (version 3.4) [42]. Effect sizes were computed using 10,000 bootstrap samples and mediation was deemed as significant if 0 was not contained within the 95% bootstrap confidence interval for an indirect effect.

### 2.6 Sample size and power analyses

A priori power calculations were conducted using G*Power (version 3.1.9.5) [43] to guide recruitment. In order to compare differences in clinical scores between DT+ and DT-populations, a minimum sample size of 86 participants per group was required to achieve 90% power, evaluated at an alpha level of 0.05 and an estimated moderate effect size (Cohen’s *d*=0.50) [44]. In order to examine the relationship between psychotic experiences and performance in the reward task, a minimum sample size of 88 participants was required to achieve 90% power evaluated at an alpha level of 0.05 and an estimated moderate critical effect size (*r*=0.3) [44].

## 3. RESULTS

### 3.1 Participant characteristics and clinical scores

Two hundred participants were recruited with 115 assigned to the DT+ group and 85 to the DT-group. Participant demographics and CTQ scores are displayed in Table 1. Age, sex, and ethnicity did not differ significantly between DT+ and DT-groups. Participants in the DT+ group were more likely to come from a lower childhood socioeconomic position, have a lower educational attainment, smoke tobacco, and were more likely to report prior access to mental health services and psychiatric medication use compared to those in the DT-group (Table 1). The DT+ group also scored significantly higher than the DT-group on measures of drug use (DAST-10), alcohol use (SMAST), anxiety (STAI), and depressive symptoms (QIDS) (Table 2). Mean CTQ scores differed between groups across all subscales (*p*<0.001; Supplementary Figure S1).

**Table 1.** Participant characteristics and demographic variables

| Sample characteristic | DT+ (*n*=115) | DT- (*n*=85) | *p* |
| --- | --- | --- | --- |
| Age (years), mean (SD) | 38.9 (12.8) | 36.9 (13.4) | 0.28<sup>a</sup> |
| Sex, *n* (%) |  |  |  |
| Female | 63 (54.8%) | 51 (60.0%) | 0.46<sup>b</sup> |
| Male | 52 (45.2%) | 34 (40%) |  |
| Ethnicity<sup>c</sup>, *n* (%) |  |  |  |
| White British | 96 (83.5%) | 65 (86.5%) | 0.48<sup>b</sup> |
| Asian | 4 (3.5%) | 7 (8.2%) |  |
| Mixed | 1 (0.9%) | 1 (1.2%) |  |
| Other | 14 (12.2%) | 12 (14.1%) |  |
| Educational attainment, *n* (%) |  |  |  |
| Degree level or above | 56 (48.7%) | 64 (75.3%) | <0.001<sup>b</sup> |
| Childhood SES<sup>d</sup>, *n* (%) |  |  |  |
| High | 38 (33.0%) | 57 (61.7%) | <0.001<sup>b</sup> |
| Intermediate | 22 (19.1%) | 13 (15.3%) |  |
| Low | 55 (47.8%) | 15 (17.6%) |  |
| Prior access to mental health services, *n* (%) | 96 (83.5%) | 41 (48.2%) | <0.001<sup>b</sup> |
| Past psychiatric medication use, *n* (%) | 61 (53.0%) | 22 (25.9%) | <0.001<sup>b</sup> |
| Tobacco smokers<sup>e</sup>, *n* (%) | 49 (42.6%) | 23 (27.1%) | 0.024<sup>b</sup> |
| CTQ score, mean (SD) | 74.9 (15.1) | 31.5 (5.0) | <0.001<sup>a</sup> |

*Abbreviations:* CTQ, Childhood Trauma Questionnaire; DAST-10, Drug Abuse Screening Test; SD, standard deviation; SES, socioeconomic status.
<sup>a</sup> Independent-samples *t*-test.
<sup>b</sup> $\chi^2$ test.
<sup>c</sup> Ethnicity was dichotomised (White British vs ethnic minority) for group comparison.
<sup>d</sup> Childhood SES was assessed by proxy measure of parental/guardian occupation (Supplementary Methods).
<sup>e</sup> Defined as a minimum of weekly tobacco smoking.

**Table 2.**
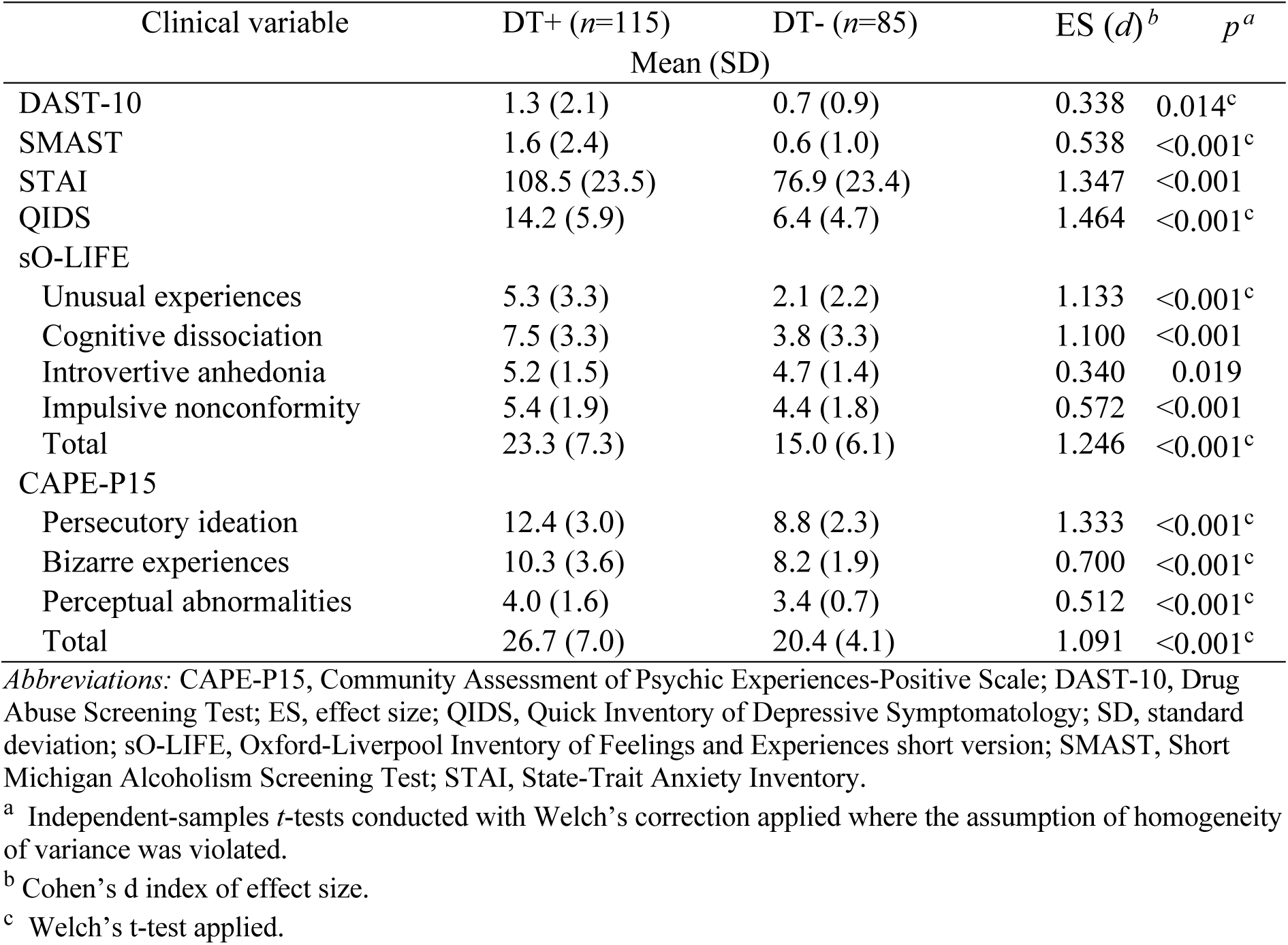
Clinical variables and subclinical scores

### 3.2 Psychotic experiences and psychosis proneness

Participants in the DT+ group had significantly higher sO-LIFE and CAPE-P15 scores across all subscales (Table 2). After adjusting for demographic variables (age, sex, ethnicity, educational attainment, childhood SES, prior access to mental health services, prior psychiatric medication use, and tobacco smoking) and clinical variables (DAST-10, SMAST, QIDS, STAI), exposure to developmental trauma predicted CAPE-P15 score (adjusted R^*2*^=0.41, R^*2*^ change=0.03, *F* change_*df*_=8.46_1,186_, *p*=0.004) and sO-LIFE score (adjusted R^*2*^=0.49, R^*2*^ change=0.03, *F* change_*df*_=7.42_1,186_, *p*=0.007). Exposure to developmental trauma was also associated with increased odds of ultra-high risk status for psychosis at the trend level (82 [71.3%] vs 19 [22.4%]; adjusted OR, 2.47; 95% CI, 0.99–6.12; *p*=0.052), adjusting for confounds. Detail of the model used, and associated findings are presented in the Supplementary Data.

### 3.3 Learning of reward contingencies

As 109 participants demonstrated below-chance behaviour, only participants who had scored ≥50% of available points were included in the analysis (DT+ *n*=50, DT-*n*=41). *Post hoc* tests revealed no difference in the proportion of DT+/DT-participants excluded (*χ*^2^ (1, *n*=91)=0.4, *p>*0.5), nor group differences in CAPE-P15 or sO-LIFE scores between participants who were included and excluded (*p*>0.6).

Analysis of acquisition performance during the probabilistic learning task using a repeated measures ANOVA showed a significant main effect of group (*F*_*df*_=6.29_1,89_, *p*=0.014), with the DT+ group demonstrating poorer task performance compared to the DT-group. A significant group × reward contingency interaction (*F*_*df*_=3.81_2,178_; *p*=0.026) was also observed. Further analysis of this interaction with planned comparisons showed that the DT+ participants were less likely to discriminate between the easiest and hardest pairs compared to controls; AB (*t*_*df*_=2.57_89_, *p*=0.012, *d*=0.54) and EF (*t*_*df*_=2.11_89_, *p*=0.014, *d*=0.45), with no significant difference in total proportion of CD choices (*t*_*df*_= -1.59_89_, *p*=0.116, *d*=0.33) (Figure 2A). These group differences in behaviour remained significant after accounting for age, sex, ethnicity, and education.

**Figure 2:**
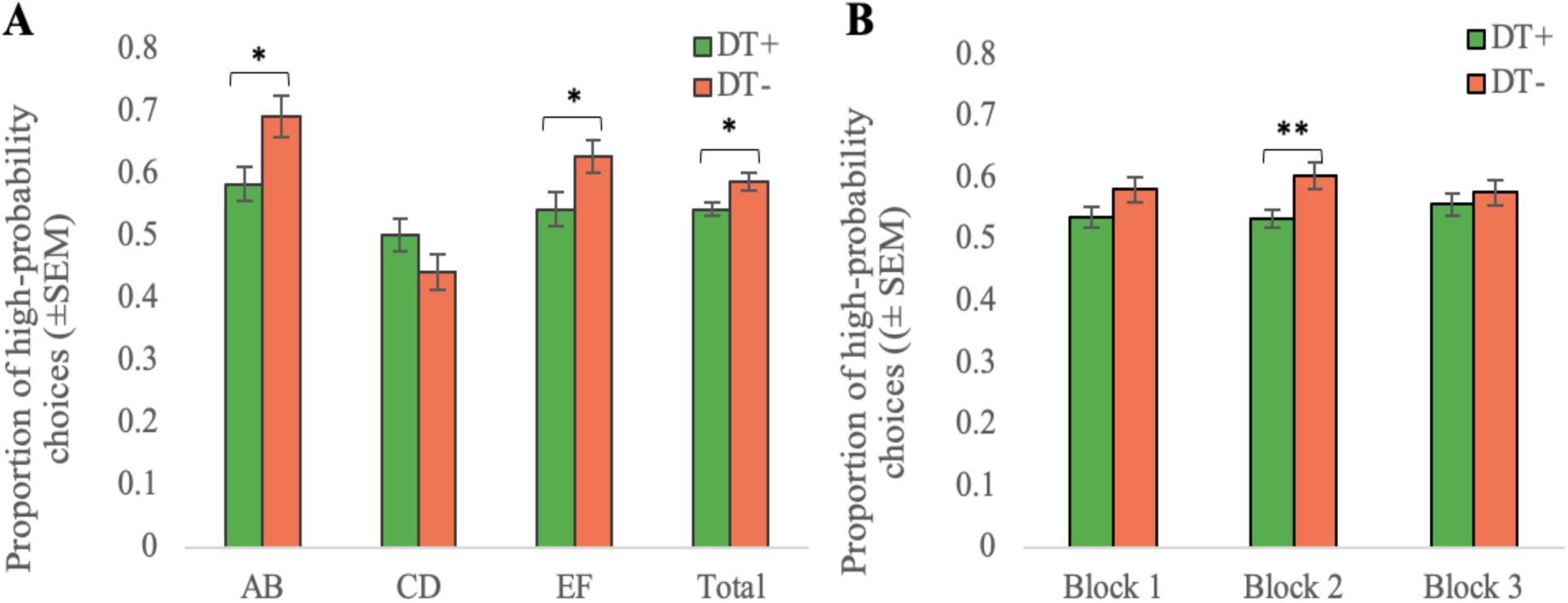
Average proportion of high-probability choices in the acquisition phase per group. (A) Stimulus-specific performance. (B) Performance by block. In both panels, error bars reflect SEM for DT+ (green) and DT-(orange) groups. \**p*<0.05, \*\**p*<0.01.

Whilst we did not observe a significant group × block interaction (*F*_*df*_=1.31_2,178_; *p*=0.275), DT+ tended to perform worse that controls in block 1 (*t*_*df*_=1.79_89_, *p*=0.077, *d*=0.37) and significantly worse in block 2 (*t*_*df*_=2.72_89_, *p*=0.008, *d*=0.56) (Figure 2B). In block 3 there was no difference in total performance between groups (*t*_*df*_=0.74_89_, *p*=0.459, *d*=0.16) (Figure 2B), suggesting that participants with exposure to developmental trauma demonstrated slower contingency learning compared to the controls. Stimulus-specific block performance is presented in Supplementary Figure 2.

### 3.4 Transfer performance

A substantial number of participants (31 out of 91 total) failed to demonstrate acquired learning of the most marked contingencies, by identifying A as the more rewarding stimulus in at least 3 out of 4 AB test trials, precluding analysis of transfer performance. As such, results from the transfer phase are reported in the Supplement.

### 3.5 Computational model

The model displayed high chain convergence to the target stationary distribution, indicated by mean Rhat values of 1.00 for all parameters, and manual examination of trace plots (Supplementary Figure S4). Model fit assessment also confirmed the Q learning gain/loss model best fit the learning phase data over compared models (Supplementary Table 2); however, loss learning rate parameters were diminished as some participants showed no appreciable learning from negative feedback (Table 3) and were therefore not analysed. DT+ participants demonstrated lower gain learning rates (Table 3; 95% HDI (−0.27, -0.01); *t*_*df*_=4.95_89_, *p*<0.001, *d*=1.01), denoting reduced learning from positive outcomes. There was no difference in exploration-exploitation parameter values across groups (95% HDI (−1.37, 0.83); *t*_*df*_=0.53_89_, *p*=0.600, *d*=0.15), indicating that group differences in learning were not due to differences in the balance between random versus deterministic approaches to trials. Supporting Bayes Factor analyses and group-level parameter posterior distributions are presented in the Supplement.

**Table 3.** Group learning parameters and highest density intervals (HDIs)

| | DT+ ($n=50$) | DT- ($n=41$) |
| --- | --- | --- |
| Parameter | Mean (95% HDI) | Mean (95% HDI) |
| $\alpha_G$ | 0.13 (0.01, 0.17) | 0.27 (0.13, 0.32) |
| $\alpha_L$ | 0.001 (0.000, 0.002) | 0.001 (0.000, 0.002) |
| $\beta$ | 1.89 (0.64, 2.49) | 2.05 (1.26, 2.26) |

### 3.5 Bivariate relationships between characterising variables and behavioural measures

There was a significant negative correlation between learning phase performance (proportion of higher-probability choices) and CAPE-P15 score (*r*= -0.26, *p*=0.034). Learning phase performance tended to be negatively associated with sO-LIFE (*r*= -0.22, *p*=0.062). Exploratory analyses revealed a trend level negative correlation between gain learning rate and CAPE-P15 score (*r*= -0.22, *p*=0.067).

### 3.6 Mediation analyses

The mediation analysis testing whether RL mediates developmental trauma in predicting psychotic experiences is shown in Figure 3. In unadjusted models, contingency learning, as measured by the total proportion of high-probability stimulus choices in the learning phase, mediated the association between developmental trauma and total CAPE-P15 score (indirect effect *β* = 0.60, 95% CI, 0.01–1.36); Figure 3), indicating that there was a significant indirect effect of developmental trauma on psychotic experiences through contingency learning.

**Figure 3:**
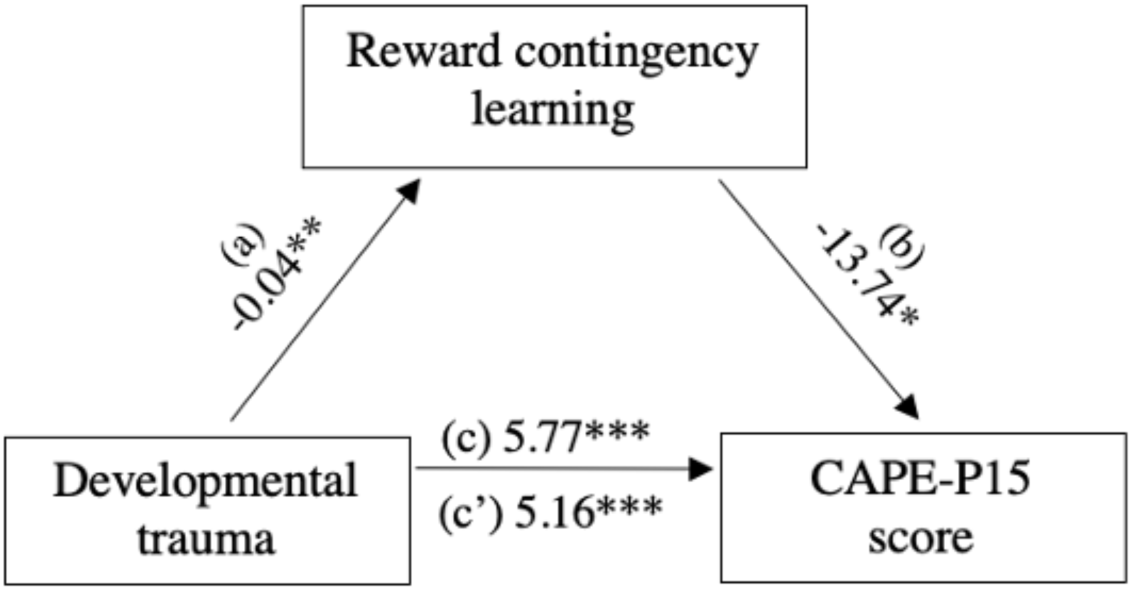
Mediation analysis. Reward contingency learning (total proportion of high-probability stimulus choices) mediated the association between developmental trauma and CAPE-P15 score indirect effect *β* = 0.60 (partially standardised *β* = 0.09), 95% CI (0.01–1.36). \**p* <0.10, \*\**p*<0.05, \*\*\**p* <0.01, unstandardised beta regression coefficients shown.

## 4. DISCUSSION

We found increased psychotic experiences and schizotypy in adult survivors of developmental trauma, compared to those without trauma histories. Adult survivors of developmental trauma demonstrated impaired learning of reward contingencies, and we found evidence that this mediated the association between developmental trauma exposure and psychotic experiences. Our results extend previous findings by showing for the first time a relationship between RL and psychotic experiences in adult survivors of developmental trauma, providing support for the hypothesis that aberrant RL is a mechanism through which exposure to developmental trauma induces vulnerability to psychosis.

### 4.1 Interpretation of results

Our finding of increased psychotic experiences and schizotypy in the developmental trauma group is consistent with the existing literature [45]. Importantly, these associations remained significant after controlling for several trauma-related covariates associated with psychotic symptoms including depression and anxiety [3,4], providing support for the notion that developmental trauma influences a detrimental shift along the psychosis continuum underlying transition to psychosis [46,47,48].

Our findings are also consistent with recent computational work demonstrating impaired RL in survivors of developmental trauma [19,20]. At the group-level, participants with developmental trauma histories demonstrated lower Bayesian-modelled learning parameters for gains, indicating that these impairments could be attributed to a reduced ability to use positive feedback to drive learning. This is in line with previous research showing developmental trauma can lead to reduced reward sensitivity [22,49,50] and attentional biases towards negative cues and threat-related stimuli [8,29] which may contribute to the impaired formation of stimulus-reward associations fundamental to RL.

The ability to detect and predict patterns in the environment (contingency detection) underpins RL and is crucial for an organism’s survival and adaptability [15]. Experiences of abuse and neglect may disrupt normative learning environments which foster the development of contingency detection. This may be especially true during development as children with experiences of developmental trauma are often subject to environmental instability and unpredictable home environments [51]. As such, survivors of developmental trauma may adaptively assume rewards are infrequent and unpredictable and apply this acquired learning in other contexts. Reduced contingency learning in survivors of developmental trauma may therefore reflect maladaptive adjustments to volatile learning environments which yield few rewards [8,20,52].

Previous studies using the probabilistic selection task in individuals with psychosis have shown that RL deficits appear to arise from impaired positive feedback-driven learning [40,53,54]. Importantly, these impairments have been linked with the severity of psychotic symptoms in these populations [53,54]. It is notable that RL deficits in our non-clinical sample of adult survivors of developmental trauma resemble those of chronically ill populations, by way of reduced learning rates following positive feedback. Our finding that deficits in RL may, in part, mediate increased psychotic experiences in individuals exposed to developmental trauma indicates that these impairments may serve as markers of latent vulnerability to psychosis. This is in line with predictive coding accounts of psychosis whereby aberrant RL contributes to inflexible belief updating through an uncorrected discrepancy between priors and perceptual information, leading to the development of hallucinations and maintenance of delusional beliefs [11,13,16,55].

The dopaminergic system is thought to play a central role in both HPC [11,18,56,57] and the pathogenesis of psychosis [58,59]. In accordance with traumatogenic neurodevelopmental and stress-diathesis models of psychosis [47,60,61], alterations to dopaminergic PE signalling, as a result of exposure to traumatic stressors, could compound the disjunction between priors and sensory information through impaired precision-weighting [56,57], making individuals with cumulative traumatic experiences increasingly prone to psychosis [48,55,62]. Indeed, dopaminergic function has been shown to be altered in survivors of developmental trauma [63,64,65] and animal models [66], and recent evidence suggests that developmental trauma may influence ventral striatal dopamine transmission to increase positive psychotic symptoms [67]. There is evidence that D_2_R antagonism may potentiate corticostriatal functional connectivity to improve reward-based RL [68]. This raises the possibility that pharmacological interventions targeting the dopaminergic system may reduce psychosis risk in trauma survivors, although further research is needed to investigate this further.

An important avenue for future research is therefore to investigate and identify the specific neurocomputational processes that underlie trauma-related alterations in RL that are associated with psychosis vulnerability. Further replication is needed to confirm these findings and it remains to be determined whether these measures predict conversion to psychosis in ultra-high risk individuals. If so, our findings suggest that RL measures may provide biomarkers to index increased risk for psychosis in individuals who have experienced developmental trauma.

### 4.2 Strengths and Limitations

A strength of this study includes the use of an experimental design with computational modelling to behaviourally probe the neurocognitive processes underlying increased psychosis risk in developmental trauma survivors. We sought to assess the combined effects of multiple forms of developmental trauma due to high rates of polyvictimisation in adult survivors [4,69].

We also examined a number of potential confounds previously linked with increased vulnerability to psychosis in trauma survivors, including comorbid anxiety and depression [69], to reduce the possibility of their confounding effects.

A limitation, however, is that our cross-sectional study design limits the ability to make causal inferences. Recall bias may also have confounded findings, although retrospective assessment of developmental trauma has been shown to be reliable in both the healthy population and patients with psychosis [31,32]. As working memory and executive function were not explicitly examined, it is possible our findings may have been reflective of a more general impairment in neurocognitive function, rather than RL per se [70]. Secondary factors including heterogeneity in the chronicity, timing, and severity of exposure, home environments during development, and subsequent stress exposure in adulthood, among other factors were not examined in this study and may have accounted for within-group differences in our sample.

Another limitation is that many participants demonstrated no appreciable learning in the acquisition of stimulus-reward contingencies, resulting in a large proportion of our sample being excluded from analysis and poor model fit of learning parameters. It is possible that participants may have been less incentivised to demonstrate optimal choice performance as the task did not offer primary rewards. To remedy this, future studies would benefit from using additional practice procedures with pre-established criteria to ascertain learning prior to completion of the transfer phase and establish reliability of our findings.

Furthermore, the task used in the study is only one example and form of RL. RL is central to understanding, interacting with and adjusting to dynamic environments. A broader assessment of RL may be more ecologically valid and improve the ability to explain the association between developmental trauma and psychosis. This may be particularly true in the context of threat processing as trauma-related impairments in RL may be associated with negative attentional and attributional biases [8,29], and aberrant safety learning [71].

In light of these caveats, future work is therefore needed to assess converging evidence at both behavioural and neural levels, resolve issues of reverse causality and elucidate specific deficits in RL which may characterise differences in the susceptibility to psychopathology.

## 5. CONCLUSION

Our study provides new insights into how psychotic symptoms may arise in adult survivors of developmental trauma. Adults survivors of developmental trauma had elevated psychotic experiences and schizotypy, and demonstrated impaired learning of reward contingencies. In line with HPC accounts of psychosis, there was evidence that aberrant RL mediated the association between developmental trauma and psychotic experiences. These findings provide supporting evidence for the hypothesis that aberrant RL is a mechanism through which developmental trauma may induce vulnerability to psychosis. Future work is therefore warranted to provide a more detailed understanding of the specific processes which characterise traumatogenic vulnerability mechanisms.

## Data Availability

The data that support the findings of this study are available from the corresponding author upon reasonable request.

## ACKNOWLEDGEMENTS

This study was funded by a UCL Excellence Fellowship to Dr Bloomfield. Dr Bloomfield is supported by the National Institute for Health Research University College London Hospitals Biomedical Research Centre.

