## Supplementary Information for "The effects of developmental trauma on reinforcement learning and its relationship to psychotic experiences: a behavioural study"

### SUPPLEMENTAL INFORMATION

#### Supplementary Materials and Methods

**Assessment of Childhood Socioeconomic Status (SES)**: Assessment of childhood SES was based on participants' self-reported highest earning parental or guardian's occupation by the time they were 18 years of age using the eight-class version of the National Statistics Socio-economic Classification (NS-SEC), refined to the three-class version, to provide measures of high (class 1), intermediate (class 2) and low (class 3) SES.

**Assessment of Developmental Trauma:** We used the self-report 25-item childhood trauma questionnaire (CTQ) [1] as a measure of developmental trauma. The CTQ measures five distinct traumatogenic domains: emotional abuse ("verbal assaults on a child's sense of worth or well-being or any humiliating or demeaning behaviour directed toward a child by an adult or older person"), physical abuse ("bodily assaults on a child by an adult or older person that posed a risk of or resulted in injury"), sexual abuse ("sexual contact or conduct between a child younger than 18 years of age and an adult or older person"), emotional neglect ("the failure of caretakers to meet children's basic emotional and psychological needs, including love, belonging, nurturance, and support"), and physical neglect ("the failure of caretakers to provide for a child's basic physical needs, including food, shelter, clothing, safety, and health care"). Scoring ranged from 1-25 for each percept. Pre-defined threshold scores [1] were used to classify exposure to each trauma type for group categorisation (presented in Table S1); at least moderate self-reported exposure to more than two types of trauma for the developmental trauma (DT+) group, and below moderate self-reported exposure to all five trauma types for the control (DT-) group.

**Table S1**. CTQ cut-off scores (as per Bernstein et al., 2003).

| | Scale | | | | | |
|---|---|---|---|---|---|---|
| Classification of exposure | Emotional abuse | Physical abuse | Sexual abuse | Emotional neglect | Physical neglect | Total score (25-125) |
| None to minimal | ≤8 | ≤7 | ≤5 | ≤9 | ≤7 | ≤36 |
| Low to moderate | 9-12 | 8-9 | 6-7 | 10-14 | 8-9 | 37-51 |
| Moderate to severe | 13-15 | 10-12 | 8-12 | 15-17 | 10-12 | 52-68 |
| Severe to extreme | ≥16 | ≥13 | ≥13 | ≥18 | ≥13 | ≥69 |

**Computational model**: The Q learning gain/loss model [2,3], an expanded version of the Rescorla-Wagner updating ('delta') rule [4], is a variation of the Q learning algorithm [5] with three free parameters; a learning rate parameters for gain outcomes ($\alpha_G$), a learning rate parameters for loss outcomes ($\alpha_L$), and an exploration-exploitation parameter ($\beta$; see below).

In the standard model, individual Q values, estimates of expected reward, are computed for each stimulus $i$ during trial $t$, and are subsequently updated on a trial-by-trial basis by the outcome $r(t)$ received: $r(t)$= 1 for reward and $r(t)$=0 for omission of reward:

$$Q_{\mathrm{i}}\,(t+1) = Q_{\mathrm{i}}(t) + \alpha_G[r(t) - Q_{\mathrm{i}}(t)]_+ + \alpha_L[r(t) - Q_{\mathrm{i}}(t)]_-$$

where the expected value of a particular stimulus ($Q_{\mathrm{i}}$) on a subsequent trial is equal to its expected value on a given trial modified by the RPE (reward received minus the reward expected), represented by $[r(t) - Q_{\mathrm{i}}(t)]$; positive or negative prediction errors are multiplied by gain and loss learning rates respectively. Q values are then computed to model stimulus-specific action selection using the 'softmax' rule:

$$P_A(t) = \frac{e^{\frac{Q_A(t)}{\beta}}}{e^{\frac{Q_A(t)}{\beta}} + e^{\frac{Q_B(t)}{\beta}}}.$$

where the exploration-exploitation parameter $\beta$, in this case, represents the probability of choosing stimulus A over stimulus B with given Q values. A higher $\beta$ value is indicative of more random choice behaviour (exploration of the available stimuli) as opposed to a lower $\beta$ value indicating more deterministic choices (selecting the stimulus believed to be more rewarding).

We implemented the Q-learning model described in a Bayesian framework (see Doll et al., 2009 for further detail) [6], and a hierarchical Bayesian approach was used to simultaneously fit group and individual parameter distributions to demonstrate acquired learning using the *hBayesDM* package for R [7]. Posterior distributions were approximated via Markov Chain Monte Carlo (MCMC) sampling; 4 MCMC chains of 4000 samples each (excluding the first 1000 as burn-in samples) were simulated and tested for adequate convergence to primary distributions by manual inspection of trace plots and Rhat value convergence to 1.00 for all parameters [8]. Higher learning rates denote increased learning from more recent outcomes, whereas lower changes in Q value on each trial are indicative of lower integrated learning over time. A confirmatory model-fit assessment was also conducted using a variation of the model (Reward-Punishment Model [9]) and a model with one learning rate parameter (the Rescorla-Wagner delta model [4]) for comparison (see Table S4).

**Hierarchical regression analyses:** We used a three-step hierarchical multiple regression model to examine the effects of developmental trauma on measures of psychotic experiences and psychosis proneness after adjusting for clinico-demographic covariates. After controlling for demographic variables in step 1 (age, sex, ethnicity, educational attainment, childhood socioeconomic status, prior access to mental health services, prior psychiatric medication use, and tobacco smoking), and clinical variables (DAST-10, SMAST, QIDS, STAI), exposure to developmental trauma (DT+ vs DT-) was entered in step 3. Dichotomised factors were dummy-coded; 0, 1 (childhood SES was coded as low= -1, intermediate= 0, high= 1). Normality of distribution and homoscedasticity were assessed using normal probability plots and residual plots. Multicollinearity was assessed using Pearson's coefficient ($r \leq 0.9$) in addition to variance inflation factor (VIF) values of <10 and tolerance values of <0.2. Independence of observations was assessed by Durbin Watson values of 1.5-2.5. All of which were not violated.

### Supplementary Data

**Developmental trauma**: As defined by more than more than 'moderate' exposure [1] (Table S1), the most common type of trauma within the DT+ group was emotional abuse (*N*=109, 95%), followed by emotional neglect (*n*=108, 94%), physical neglect (*n*=93, 81%), physical abuse (*n*=72, 63%), and sexual abuse (*n*= 56, 49%). 37% of participants (*n*=43) reported three types of trauma, 44% (*n*=51) reported four types, and 18% (*n*=21) reported all five CTQ subtypes. Mean CTQ subscale scores for the DT+ and DT- groups are presented below.

**CTQ scores**

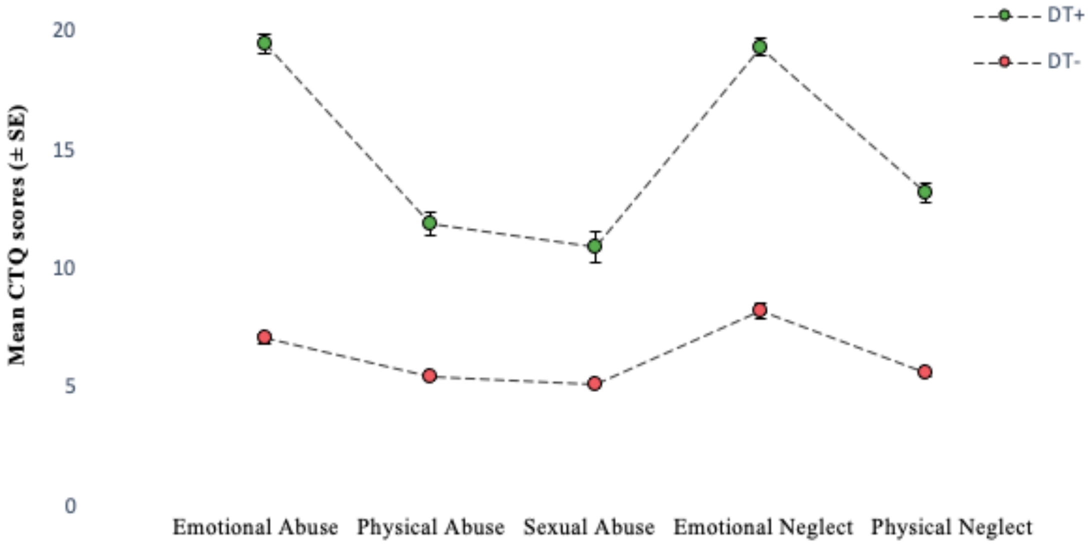


**Figure S1**: Mean CTQ subscale scores for DT+ (green) and DT- (orange) groups. Error bars reflect SEM. Score cut-offs are presented in Table S1.

**Table S2.** Hierarchical regression model for variables predicting total CAPE-P15 score.

| **Outcome variable: CAPE-P15 – total** | | | | |
|---|---|---|---|---|
| | *Unstandardised B* | *B* 95% CI (lower, upper) | *Standardised β* | *p* |
| Model 1[a] | | | | |
| Constant | 28.21 | 24.63, 31.79 | | 0.000 |
| Age | -0.11 | -0.19, -0.04 | -0.22 | 0.004 |
| Sex | -1.30 | -3.17, 0.56 | -0.10 | 0.169 |
| Ethnicity | -0.08 | -2.44, 2.28 | -0.01 | 0.945 |
| Education | -2.30 | -4.23, -0.37 | -0.17 | 0.020 |
| Childhood SES | 0.27 | -0.77, 1.30 | 0.04 | 0.611 |
| Smoking | 1.03 | -0.93, 2.99 | 0.08 | 0.300 |
| Prior access of mental health services | 1.54 | -0.737, 3.81 | 0.11 | 0.184 |
| Past medication use | 1.83 | -0.33, 3.98 | 0.14 | 0.097 |
| Model 2[b] | | | | |
| Constant | 15.31 | 10.80, 19.83 | | 0.000 |
| Age | -0.04 | -0.11, 0.02 | -0.08 | 0.186 |
| Sex | -0.16 | -1.77, 1.44 | -0.01 | 0.841 |
| Ethnicity | 0.69 | -1.26, 2.65 | 0.04 | 0.484 |
| Education | -1.41 | -3.06, 0.23 | -0.11 | 0.092 |
| Childhood SES | 0.50 | -0.33, 1.39 | 0.07 | 0.226 |
| Smoking | 0.050 | -1.62, 1.72 | 0.00 | 0.952 |
| Prior access of mental health services | -0.97 | -2.93, 0.98 | -0.07 | 0.328 |
| Past medication use | 0.21 | -1.61, 2.03 | 0.02 | 0.819 |
| DAST-10 | 0.54 | 0.06, 1.03 | 0.14 | 0.029 |
| SMAST | 0.16 | -0.26, 0.57 | 0.05 | 0.455 |
| QIDS | 0.25 | 0.05, 0.45 | 0.25 | 0.015 |
| STAI | 0.08 | 0.03, 0.13 | 0.35 | 0.001 |
| Model 3[c] | | | | |
| Constant | 16.10 | 11.64, 20.57 | | 0.000 |
| Age | -0.05 | -0.12, 0.01 | -0.10 | 0.098 |
| Sex | 0.02 | -1.56, 1.60 | 0.002 | 0.976 |
| Ethnicity | 0.57 | -1.35, 2.48 | 0.03 | 0.561 |
| Education | -1.16 | -2.78, 0.47 | -0.09 | 0.162 |
| Childhood SES | 0.96 | 0.07, 1.85 | 0.13 | 0.036 |
| Smoking | -0.12 | -1.77, 1.52 | -0.01 | 0.883 |
| Prior access of mental health services | -1.32 | -3.25, 0.61 | -0.09 | 0.179 |
| Past medication use | 0.18 | -1.60, 1.96 | 0.01 | 0.845 |
| DAST-10 | 0.58 | 0.10, 1.05 | 0.15 | 0.019 |
| SMAST | 0.11 | -0.29, 0.52 | 0.03 | 0.582 |
| QIDS | 0.19 | -0.01, 0.39 | 0.19 | 0.066 |
| STAI | 0.07 | 0.02, 0.11 | 0.29 | 0.005 |
| Developmental trauma | 2.30 | 0.97, 5.03 | 0.22 | 0.004 |

[a] $R^2$ = 0.13, adjusted $R^2$ = 0.09, $F_{df}$= $3.551_{8,191}$, $p$=0.001

[b] $\Delta R^2$ = 0.29, adjusted $R^2$ = 0.39, $F_{df}$= $23.637_{4,187}$, $p$<0.001

[c] $\Delta R^2$ = 0.03, adjusted $R^2$ = 0.41, $F_{df}$= $8.462_{1,186}$, $p$=0.004

**Table S3.** Hierarchical regression model for variables predicting total sO-LIFE score.

| **Outcome variable: sO-LIFE – total** | | | | |
|---|---|---|---|---|
| | *Unstandardised B* | *B* 95% CI (lower, upper) | *Standardised β* | *p* |
| Model 1[a] | | | | |
| Constant | 21.94 | 17.67, 26.21 | | 0.000 |
| Age | -0.07 | -0.16, 0.02 | -0.12 | 0.112 |
| Sex | -2.76 | -4.98, -0.54 | -0.17 | 0.015 |
| Ethnicity | 0.41 | -2.41, 3.22 | 0.02 | 0.776 |
| Education | -2.10 | -4.40, 0.20 | -0.13 | 0.074 |
| Childhood SES | -0.41 | -1.64, 0.82 | -0.05 | 0.507 |
| Smoking | 1.37 | -0.97, 3.70 | 0.08 | 0.251 |
| Prior access of mental health services | 2.52 | -0.19, 5.23 | 0.15 | 0.068 |
| Past medication use | 2.22 | -0.36, 4.79 | 0.14 | 0.091 |
| Model 2[b] | | | | |
| Constant | 3.15 | -1.87, 8.17 | | 0.218 |
| Age | 0.02 | -0.05, 0.09 | 0.04 | 0.503 |
| Sex | -1.29 | -3.08, 0.49 | -0.08 | 0.155 |
| Ethnicity | 1.49 | -0.68, 3.66 | 0.07 | 0.177 |
| Education | -1.27 | -3.10, 0.56 | -0.08 | 0.174 |
| Childhood SES | -0.02 | -0.97, 0.94 | -0.002 | 0.974 |
| Smoking | 0.17 | -1.69. 2.03 | 0.01 | 0.859 |
| Prior access of mental health services | -0.91 | -3.08, 1.26 | -0.05 | 0.408 |
| Past medication use | 0.21 | -1.81, 2.22 | 0.01 | 0.840 |
| DAST-10 | 0.78 | 0.24, 1.32 | 0.17 | 0.005 |
| SMAST | -0.05 | -0.51, 0.41 | -0.01 | 0.831 |
| QIDS | 0.21 | -0.02, 0.43 | 0.17 | 0.069 |
| STAI | 0.14 | 0.09, 0.19 | 0.50 | 0.000 |
| Model 3[c] | | | | |
| Constant | 3.97 | -1.00, 8.94 | | 0.117 |
| Age | 0.01 | -0.06, 0.08 | 0.02 | 0.703 |
| Sex | -1.10 | -2.86, 0.66 | -0.07 | 0.220 |
| Ethnicity | 1.36 | -0.78, 3.50 | 0.07 | 0.211 |
| Education | -1.00 | -2.81, 0.81 | -0.06 | 0.279 |
| Childhood SES | 0.43 | -0.56, 1.42 | 0.05 | 0.394 |
| Smoking | -0.01 | -1.85, 1.82 | -0.001 | 0.989 |
| Prior access of mental health services | -1.28 | -3.43, 0.87 | -0.08 | 0.243 |
| Past medication use | 0.17 | -1.81, 2.16 | 0.01 | 0.866 |
| DAST-10 | 0.81 | 0.28, 1.35 | 0.18 | 0.003 |
| SMAST | -0.09 | -0.55, 0.36 | -0.02 | 0.680 |
| QIDS | 0.14 | -0.08, 0.36 | 0.12 | 0.212 |
| STAI | 0.13 | 0.08, 0.18 | 0.45 | 0.000 |
| Developmental trauma | 3.13 | 0.86, 5.39 | 0.20 | 0.007 |

[a] $R^2 = 0.14$, adjusted $R^2 = 0.10$, $F_{df}$= $3.80_{8,191}$, $p$=<0.001

[b] $\Delta R^2 = 0.37$, adjusted $R^2 = 0.47$, $F_{df}$= $34.36_{4,187}$, $p$<0.001

[c] $\Delta R^2 = 0.02$, adjusted $R^2 = 0.49$, $F_{df}$= $7.42_{1,186}$, $p$=0.007

**Stimulus-specific performance per block**

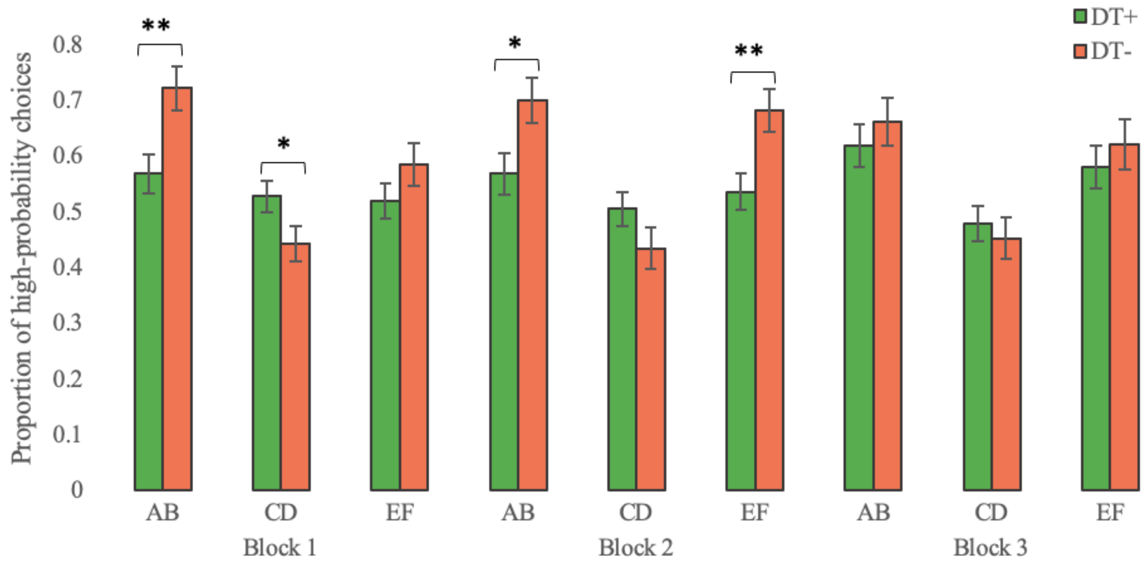


**Figure S2:** Average proportion of high-probability choices for each stimulus pair within each block. Error bars reflect SEM for DT+ (green) and DT- (orange) groups. *$p$ <0.05, **$p$<0.01.

**Transfer performance**: after initial learning, analysis of transfer phase performance revealed no significant group difference between DT+ and DT- groups in positive feedback (Go) learning, as indicated by choose-A choices, nor negative feedback (NoGo) learning, as indicated by avoid-B choices. Group differences remained non-significant when excluding subjects who failed to demonstrate learning of the AB contingencies by identifying A as the most rewarding stimulus in at least 3 out of 4 AB test trials ($t_{df}$=0.48$_{58}$, $p$=0.636; $t_{df}$=1.41$_{89}$, $p$=0.165) (Figure S3).

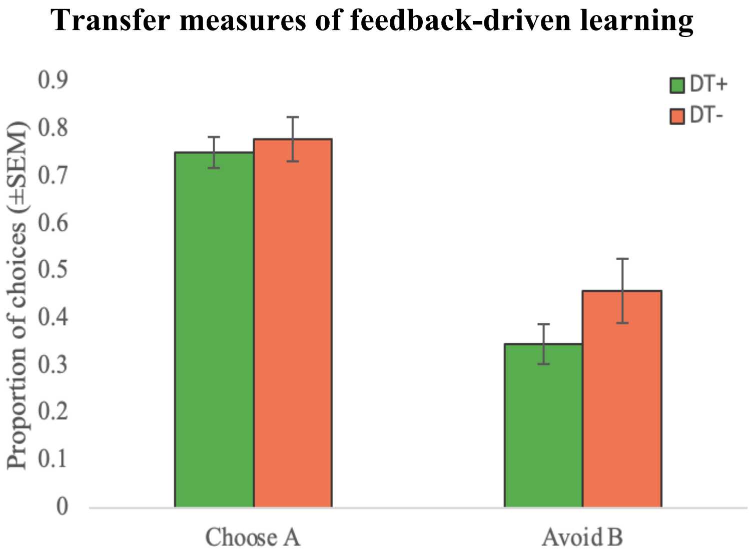


**Figure S3:** Proportion of choose-A and avoid-B choices in the transfer phase. Transfer measures of positive feedback-driven learning was assessed by choosing the most rewarded stimulus in test trials (choose-A) and negative feedback-driven learning by avoiding the least rewarded stimulus (avoid-B). Error bars reflect SEM for DT+ (green; *n*=30) and DT- (orange; *n*=30) groups.

**Table S4.** Computational model fit assessment for each group using the leave-one-out cross-validation information criterion (LOOIC) procedure.

| Model | LOOIC($_{DT+}$) | LOOIC($_{DT-}$) | LOOIC($_{Sum}$) |
|---|---|---|---|
| PST-Q learning[a] | 11258.5 | 8874.1 | 20132.6 |
| RWD | 12267.0 | 9711.7 | 21978.8 |
| PRL-RP | 11742.3 | 8874.7 | 20617.0 |

*Abbreviations:* LOOIC, leave-one-out cross-validation information criterion; PRL-RP, probabilistic reversal learning task reward-punishment model; PST-Q learning, probabilistic selection task gain/loss Q learning model; RWD, Rescorla-Wagner delta model.

[a] The gain/loss Q learning model (Frank et al., 2007) was found to be the best fitting model compared to the Rescorla-Wagner delta model (Rescorla and Wagner, 1972) and Reward-Punishment model (den Ouden et al., 2013), as indicated by lower LOOIC values.

### Markov chain trace plots for separately modelled group parameters

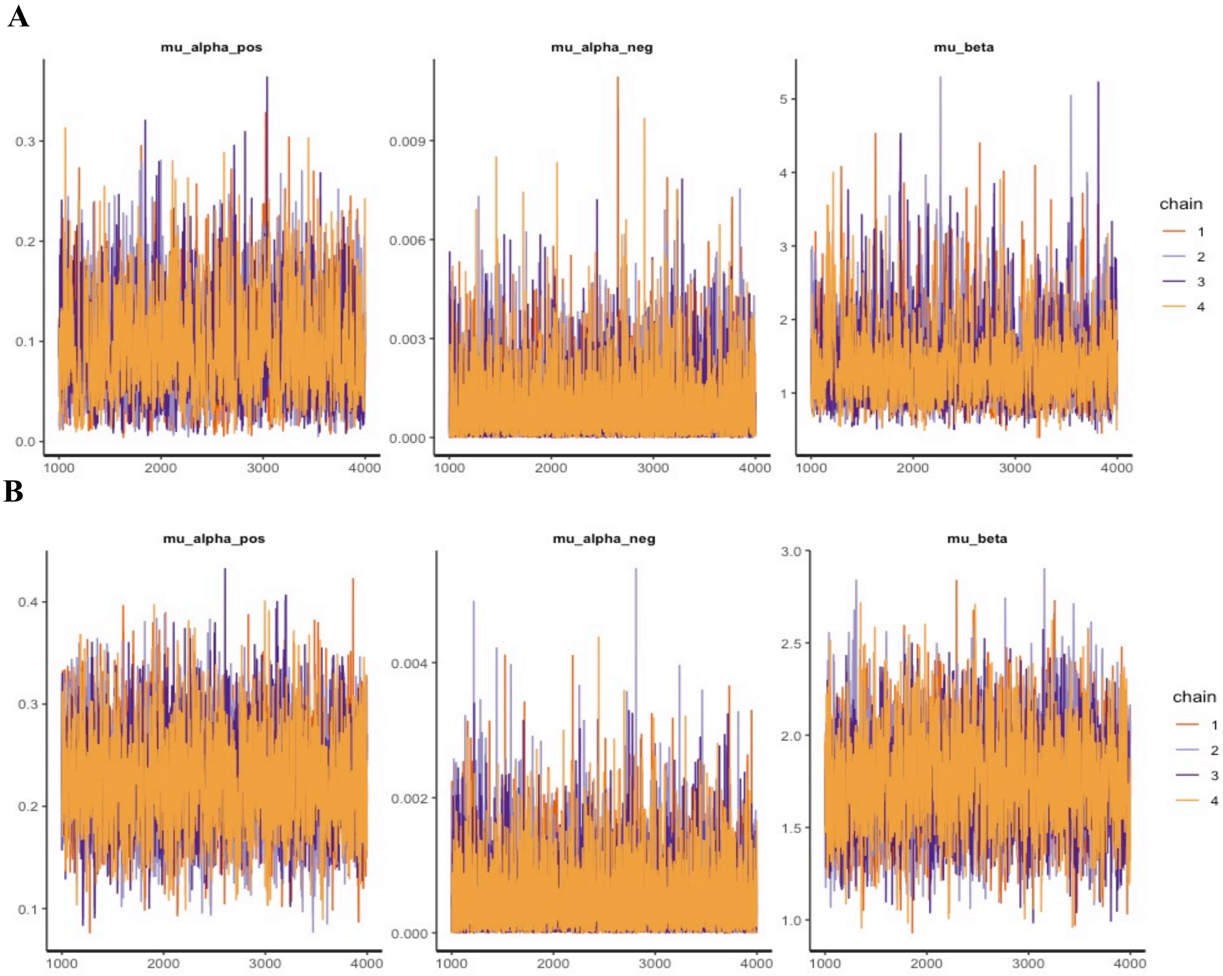


**Figure S4**: Trace plots showing approximated parameter vectors for DT+ (A) and DT- (B) groups run with over 4000 iterations of 4 Markov chains, excluding 1000 warm-up samples.

**Table S5.** Group learning parameters and highest density intervals

| | DT+ (*n*=50) | DT- (*n*=41) | DT+ vs DT- | | |
|---|---|---|---|---|---|
| Parameter | Mean (95% HDI) | Mean (95% HDI) | 95% HDI | BF [a] | *p* [b] |
| $\alpha_G$ | 0.13 (0.01, 0.17) | 0.27 (0.13, 0.32) | -0.27, -0.01 | 4852.31[c] | *p*<0.001 |
| $\alpha_L$ | 0.001 (0.000, 0.002) | 0.001 (0.000, 0.002) | -0.002, 0.003 | - | - |
| $\beta$ | 1.89 (0.64, 2.49) | 2.05 (1.26, 2.26) | -1.37, 0.83 | 4.011[d] | *p*=0.600 |

[a] Bayes Factor analyses using the Jeffreys-Zellner-Siow prior (*JZS*) with a scale factor of 0.707. Bayes factors of >10 indicate strong evidence for a difference, Bayes factors of >100 indicate decisive evidence for a difference as per Harold Jeffreys interpretation.

[b] Frequentist analyses (independent *t*-tests) on means of compared posterior distributions.

[c] Significant evidence for lower $\alpha_G$ positive learning in the DT+ group compared to the DT- group was found (BF>100).

[d] No significant difference in exploitation-exploration was found between groups (BF= 4.01).

### Group level differences in Q-learning model parameters

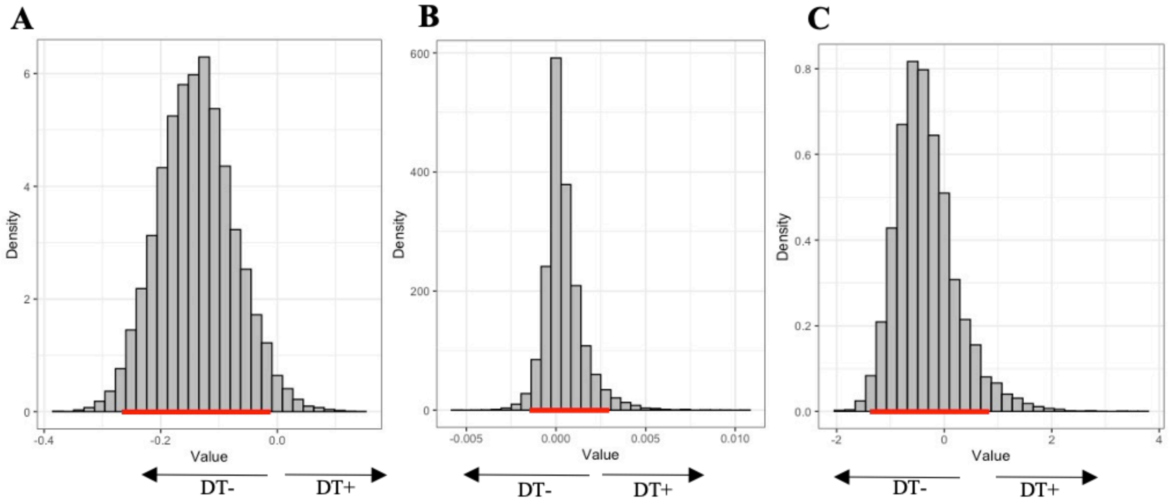


**Figure S5**: Highest density interval (HDI) plots for group level differences in parameters. (A) $\alpha_G$ parameter (95% HDI: -0.27, -0.01). The large leftward shift indicates higher $\alpha_G$ values in the DT- group. (B) $\alpha_L$ parameter (95% HDI: -0.002, 0.003). (C) $\beta$ parameter (95% HDI: -1.37, 0.83).
